# Challenging mtDNA tRNA Variant Guidelines: Emphasizing Single-Cell Analysis through Four Novel Variants

**DOI:** 10.1101/2025.11.19.25339649

**Authors:** Prosper Chloé, Zereg Elamine, Chaussenot Annabelle, Bannwarth Sylvie, Lannes Béatrice, Nathalie Streichenberger, Kaphan Elsa, Nadaj-Pakleza Aleksandra, Villa Luisa, Sacconi Sabrina, Masingue Marion, Francou Bruno, Ait-El-Mkadem Saadi Samira, Fragaki Konstantina, Paquis-Flucklinger Véronique, Rouzier Cécile

**Author notes:** **Correspondence** Dr Cécile Rouzier, Service de Génétique Médicale, Laboratoire de Biologie Moléculaire, Centre de Référence des Maladies Mitochondriales, Hôpital L’Archet2, 151 Route de Saint Antoine de Ginestière, 06200, Nice, France.

## Abstract

The broad clinical and genetic heterogeneity of mitochondrial diseases makes diagnosis challenging. Accurate characterization of novel variants is crucial to reduce diagnostic uncertainty, guide treatment, and enable reliable genetic counseling. In this study, we validated a single-cell NGS-based analysis approach by comparison with conventional PCR-RFLP and applied it to five mtDNA VUS identified in patients evaluated at our national reference center for mitochondrial disorders (CALISSON). Variant interpretation was assessed using multiple frameworks, including Yarham’s scoring, Wong’s specifications, and the ClinGen guidelines, highlighting differences between these scoring systems and the limitations of current recommendations in fully integrating functional evidence and tissue segregation data. We implemented a classification approach that incorporates these aspects to achieve a more clinically meaningful interpretation. This analysis enabled the reclassification of four novel variants (m.9998T>C in *MT-TG*, m.7530A>G in *MT-TD*, and m.4271G>C and m.4305A>G in *MT-TI*), providing a definitive molecular diagnosis. Further validation on a larger set of variants will be required, as well as the establishment of standardized criteria for single-fiber analyses, including minimum fiber numbers, thresholds for COX-negative fibers, and statistical significance. Overall, this study underscores the critical importance of integrating robust functional evidence into mtDNA variant interpretation and provides insights for refining existing guidelines to improve diagnostic accuracy and support clinical decision-making.

## 1. Introduction

Mitochondrial diseases result from defects in the mitochondrial respiratory chain, caused by genetic variants that impair oxidative phosphorylation. These variants are located either in mitochondrial DNA (mtDNA) or nuclear genes^1^ leading to a mix of Mendelian and maternal inheritance patterns^2,3^. When mitochondrial disease is suspected, a comprehensive analysis of mtDNA through Next Generation Sequencing (NGS) is an essential first step^1^. The diagnostic yield of mtDNA sequencing is estimated to be approximately 10-15%, depending on the clinical indication, with approximately 10% of detected variants classified as variants of uncertain significance (VUS).

Clarifying the pathogenicity of mitochondrial VUS is essential for accurate diagnosis and informed patient management. While the classification of mtDNA variants follows principles similar to those applied to nuclear DNA, it requires additional criteria that account for the unique features of the mitochondrial genome, particularly heteroplasmy. Most pathogenic mtDNA variants are heteroplasmic, with mutant and wild-type genomes coexisting within the same cell or tissue. The heteroplasmy level can vary significantly between tissues and plays a critical role in determining disease severity and phenotypic expression, due to the existence of a threshold effect^4,5^. Furthermore, mtDNA mutations frequently involve tRNA genes, and assigning pathogenicity to these variants can be particularly challenging. Early frameworks proposed by McFarland et al. and Yarham et al. introduced comprehensive scoring systems to evaluate the pathogenic potential of mitochondrial tRNA variants^6,7^. More recently, following the publication of the ACMG/AMP guidelines to harmonize variant interpretation practices, it became necessary to adapt these criteria to the mitochondrial genome, first through the specifications proposed by Wong et al. and later refined by the ClinGen working group. Regarding functional validation criteria (PS3), experimental evidence is primarily obtained from cybrid studies and single-fiber analyses^8–10^.

The single-fiber analysis quantifies heteroplasmy levels in individual muscle fibers and correlates these with biochemical deficiencies, such as cytochrome c oxidase (COX) activity^11^. In a previous study conducted by Zereg et al.^12^, we implemented a routine strategy for single-fiber analysis using fluorescence PCR–restriction fragment length polymorphism (PCR-RFLP) to quantify the heteroplasmy levels of mtDNA variants, which enabled us to confirm the molecular diagnosis in six patients. Furthermore, we addressed the challenge of interpreting pathogenic variants identified at low levels of heteroplasmy, demonstrating in one patient the contribution of single-fiber studies in such cases. This method improved our laboratory’s diagnostic yield and patient management. However, PCR-RFLP is a time-consuming technique that requires specific technical design for each variant and remains challenging to sustain in routine diagnostic workflows. Considering the large and growing number of potentially pathogenic mtDNA variants, we employed an NGS-based approach in this study that simplifies the design process and enables accurate quantification of heteroplasmy levels. First, we validated this approach by comparing it with the conventional PCR-RFLP technique. We then applied it to five mtDNA VUS identified in patients tested at our reference center for mitochondrial disorders (CALISSON). The interpretation of these variants was subsequently assessed using several existing guidelines. Despite the use of single-cell analysis, considered the gold standard for functional validation, we found that recent recommendations often did not allow these variants to be reclassified as pathogenic. We therefore critically reviewed the current criteria and proposed adaptations for future versions of these guidelines that would assign greater weight to robust functional evidence.

## 2. Materials and methods

### 2.1 Patients

We investigated five patients presenting clinical features suggestive of mitochondrial dysfunction and cytochrome c oxidase (COX)-deficient fibers on histological examination of muscle biopsy (Table 1). Biochemical analyses, as well as mtDNA (GenBank number NC_012920) sequencing on muscle, urine, or blood included in the routine strategy, were performed as previously described^12^. For each patient, a variant affecting a tRNA and classified as a VUS at the time of sequencing had been identified (Table 1, Fig 1).

**Fig 1.**
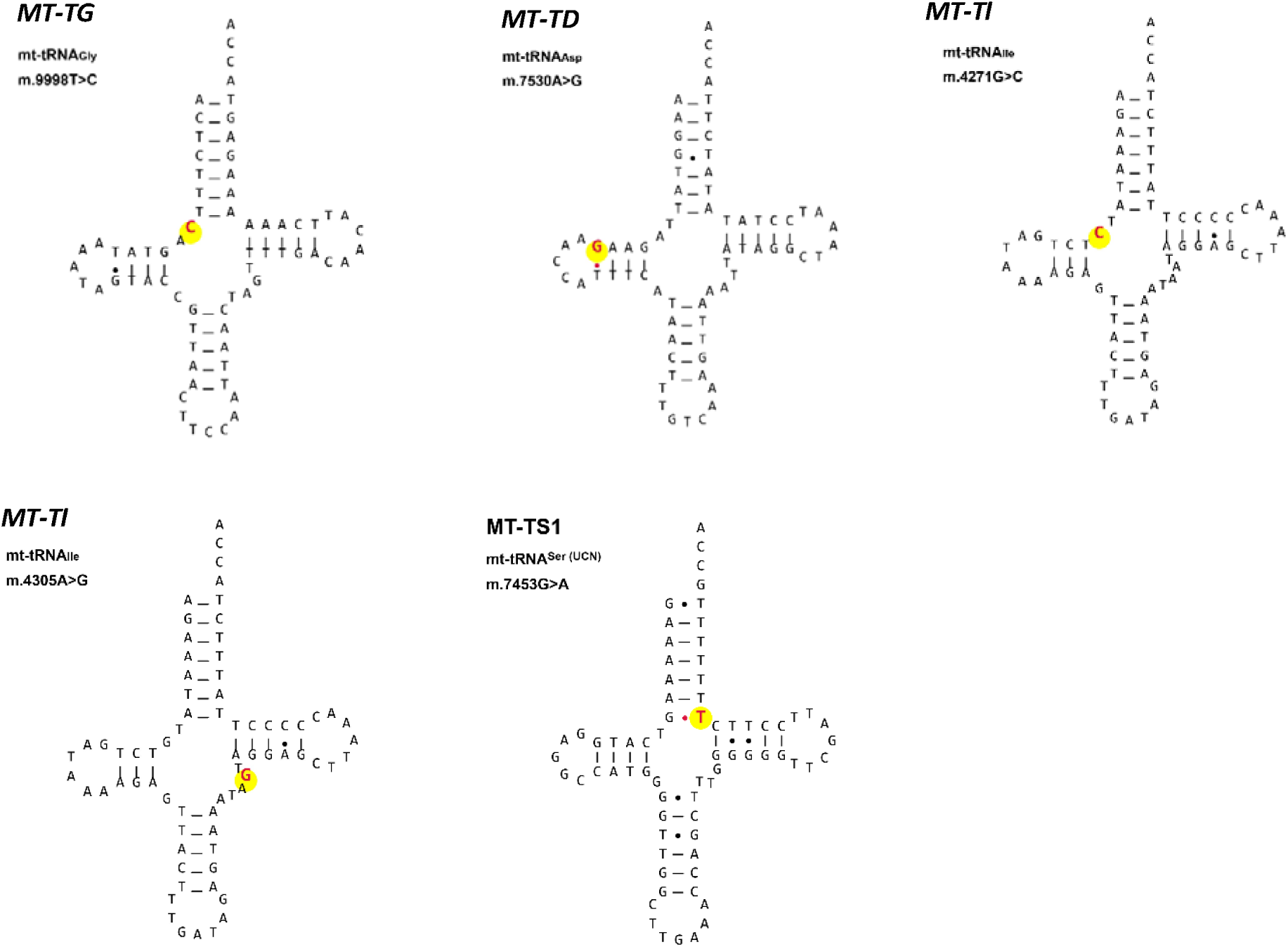
mt-tRNA secondary structures demonstrating the location of the five mt-tRNA variants identified. The location of each mt-tRNA variant within the tRNA cloverleaf structure is shown, using MitoVizualize ^17^. mt-tRNAs = mitochondrial transfer RNAs

**Table 1.** Overview of patient clinical, biochemical and genetic data.

| Patient details<sup>a</sup> | Clinical presentation | Muscle histology<sup>b</sup> | Respiratory chain deficiency<sup>c</sup> | Mt-DNA variant and location | In silico prediction<sup>d</sup> | Conservation<sup>e</sup> | Mt-DNA deletion<sup>f</sup> | % mutation load in patient tissues<sup>g</sup> | % mutation load in familial tissues<sup>h</sup> | Previous reports |
| --- | --- | --- | --- | --- | --- | --- | --- | --- | --- | --- |
| **1**<br>(M, 35-40) | Exercise intolerance, cataract, optic neuropathy, deafness, growth delay, hypogonadism, diabetes insipidus, behavioral & cognitive disorder, recurrent coma with lactic acidosis, epilepsy & status epilepticus (death 35-40 years old)<br>MRI : mild T2/FLAIR hypersignals of basal ganglia | RRF +, 80% COX- | III, IV | m.9998T>C (tRNA Gly, MT-TG) | 11.63 (40%)<br>Possibly benign | 100% | N | M = 97%<br>B = 21%<br>U = 76.6% | Unaffected mother<br>U = 0%<br>B = 0%<br>BM = 0% | none |
| **2**<br>(F, 35-40) | Deafness, myoclony, cognitive disorders, dystonia, spasticity, ptosis, cerebellar syndrome, cataract, RP | RRF +, 50% COX- | IV | m.7530A>G (tRNA Asp, MT-TD) | 17.75 (>75%)<br>Likely pathogenic | 100 % | N | M = 95%<br>B = 0 %<br>BM = 32% | Non available | none |
| **3**<br>(M, 30-35) | Deafness, RP, epilepsy, growth delay | RRF +, 60% COX- | I-III-IV | m.4305A>G (tRNA Ile, MT-TI) | 12.25 (44.5%)<br>Possibly benign | 97.78% | N | M = 76%<br>B = 21% | Non available | none |
| **4**<br>(F, 70-75) | Exercise intolerance, myalgia, proximal muscular weakness | 1 RRF, lipid overload, few COX- | N | m.4271G>C (tRNA Ile, MT-TI) | 12.87 (52.2%)<br>Possibly pathogenic | 100% | Multiple deletions | M = 16%<br>B = 4%<br>U = 6% | Non available | none |
| **5**<br>(M, 40-45) | Initial episode of elevated CPK, ptosis and diplopia myalgia, exercise intolerance, fatigue, proximal and distal weakness of 4 limb, hearing loss, cataract, gastrointestinal dysmotility | Many RRF, lipid overload, 30% COX- | N | m.7453G>A (tRNA Ser, MT-TS1) | 15.171 (68%)<br>Possibly pathogenic | 93.33% | N | M = 96%<br>B = 35%<br>U = 43% | Non available | <sup>13</sup><br><sup>14</sup><br><sup>15</sup> |

For all patients, informed consent was obtained during the diagnostic process. Additional consent was obtained as part of a research project (16-AOIP-04 [sponsor: CHU de Nice], IDRCB No.: 2017- A00688-45).

### 2.2 Single muscle fiber isolation and sample preparation

Skeletal muscle cryosections (20μm) were COX/SDH stained on 1mm PEN-membrane microscope slides (Zeiss). We used laser capture microdissection of COX-positive and COX negative fibers, followed by PCR reaction, as previously described^12^. We dissected at least five COX-positive and five COX-negative fibers for each patient.

### 2.3 Assessment of mtDNA heteroplasmy

#### NGS

After overnight incubation of each single-fiber sample, 15 µL was used in a standard 50 µL PCR reaction. PCR amplicons were obtained using primers listed in Table S1 and 2.5 U AmpliTaq polymerase. PCR conditions included an initial denaturation at 95°C for 15 min, followed by 40 cycles at 95°C for 15 s, 54–60°C for 15 s, and elongation at 72°C for 30 s. A final elongation step was performed at 72°C for 5 min (Table S1). PCR amplicons were purified using AMPure XP and 70% ethanol. Sequencing of purified PCR amplicons was performed as previously described^18^ using the Ion GeneStudio S5.

#### PCR-RFLP

The procedure was performed as described by Zereg et al.^12^. PCR amplicons were digested with the appropriate restriction enzymes, and mutant load was quantified by calculating the relative peak areas, with a sensitivity threshold of 5%.

### 2.4 Assigning pathogenicity to mitochondrial variants

To compare the different guidelines for mtDNA variant classification, the pathogenicity of each mt-tRNA variant identified in this study was assessed using three previously published approaches: the scoring system established by Yarham et al.^6^, and two adaptations of the ACMG guidelines^8^, one published by Wong et al. specifically designed for tRNA variants^9^ and the most recent by the ClinGen Expert Panel^10^.

### 2.5 Statistical analysis

The comparison of heteroplasmy levels in single cells obtained by PCR-RFLP and NGS was performed using the U Mann-Whitney test with StatView software (SAS Institute, Inc., Cary, NC). A p-value > 0.05 was considered to indicate no significant difference between the groups.

Differences in heteroplasmy levels between COX+ and COX− cells were analyzed using the unpaired t-test with StatView software (SAS Institute, Inc., Cary, NC). Values were considered significant at p < 0.05.

## 3. Results

### 3.1 Validation of heteroplasmy quantification by NGS

Using two variants from two patients previously reported by Zereg et al.^12^, we compared heteroplasmy levels in single muscle fibers using both NGS and PCR-RFLP. Heteroplasmy quantification was performed on 20 muscle fibers (5 COX-deficient and 5 COX-positive fibers from each patient), with both methods applied to each fiber. Statistical analysis using the U Mann-Whitney test revealed no significant difference between the two techniques (U = 288.0, p = 0.613). These findings indicate that NGS and PCR-RFLP provide statistically comparable heteroplasmy measurements (Fig. 2).

**Fig 2.**
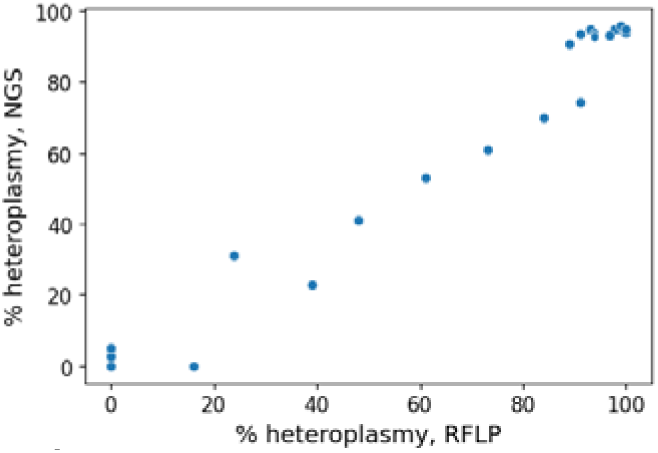
Comparison of NGS and PCR-RFLP techniques.

### 3.2 Assessment of mtDNA heteroplasmy in single cells using NGS

For each variant, m.9998T>C, m.7530A>G, m.4305A>G, m.4271G>C and m.7453G>A, we performed single-cell heteroplasmy quantification using NGS data to assess whether the mutation rate in individual COX-positive and COX-negative fibers correlated with the observed biochemical phenotype. Significantly higher heteroplasmy levels (p < 0.05) were observed in COX-negative fibers compared to COX-positive fibers in all five patients (Fig. 3). For all variants except m.4271G>C (patient 4), heteroplasmy levels in COX-negative fibers were above 95%.

**Fig 3.**
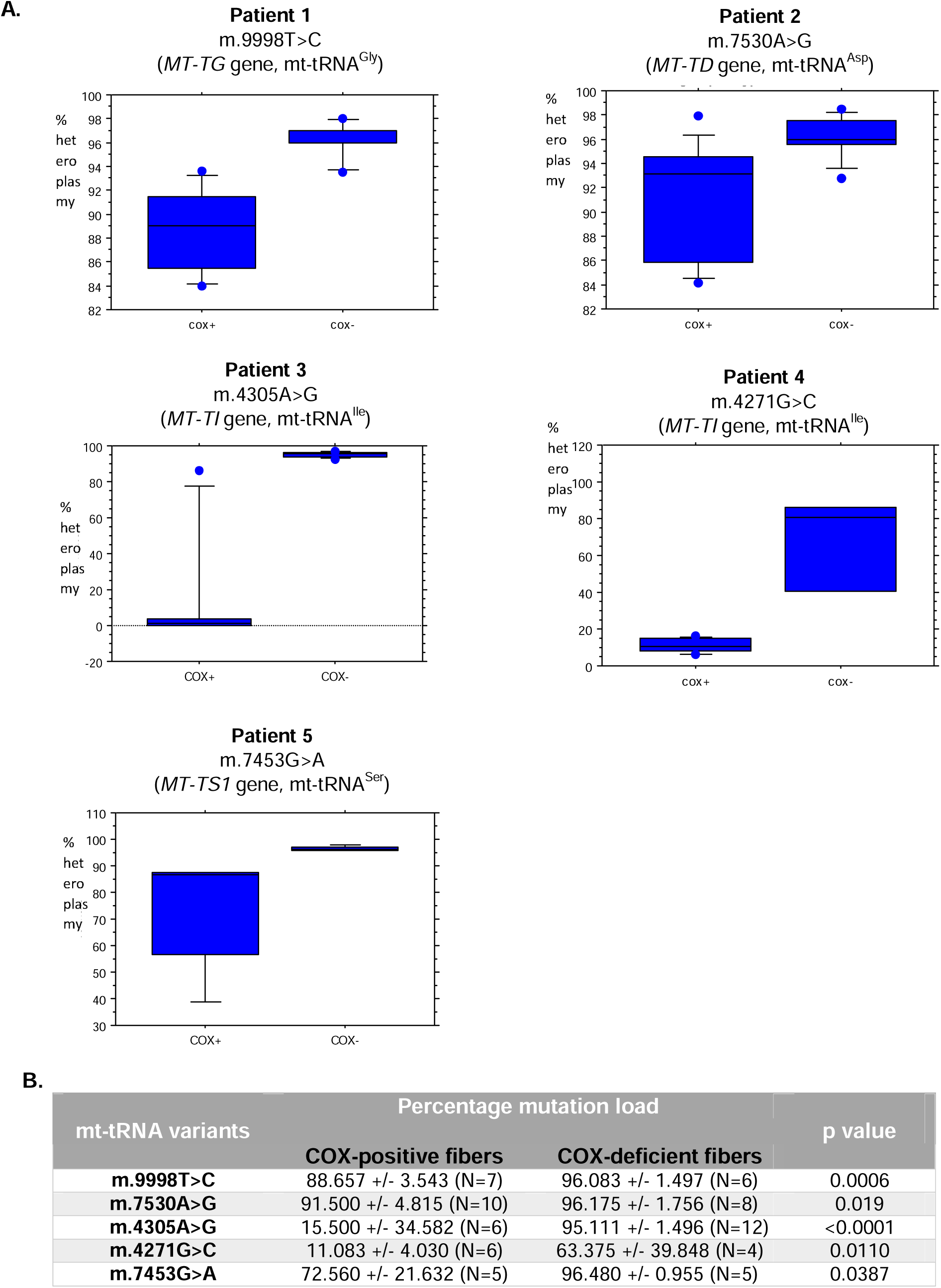
Heteroplasmy levels in single cells obtained by NGS. **A.** Boxplots showing single cell heteroplasmy quantification of the five mtDNA variants in COX+ and COX- fibers. **B**. Data of the single-fiber mutation load. COX = cytochrome c oxidase; mt-tRNA= mitochondrial transfer RNA

### 3.3 Classification of mt-tRNA variants

The classification of the five mtDNA variants according to the three guidelines^6,9,10^, before and after performing the single-fiber analysis, is presented in Table 2. Although at the time of identification, the m.7453G>A variant had been considered a VUS due to insufficient evidence, a more recent study^13^ confirmed its pathogenicity, which was taken into account in the classification presented here. However, because the clinical phenotype observed in patient 5 differs significantly from that previously reported, we included this variant in the present study.

**Table 2.** Classification of mt-tRNA variants according to the three previous guidelines as well as our suggested classification before and after inclusion of single fiber analysis. (PM7 = PP3_moderate, PM10 = PS3_moderate, PP7 = PM2_supporting)

| *mt-tRNA variants* | *Yarham et al.*<br>*(before SF study)* | *Yarham et al.* | *Wong et al.*<br>*(before SF study)* | *Wong et al.* | *ClinGen*<br>*(before SF study)* | *ClinGen* | *Suggested scoring*<br>*(before SF study)* | *Suggested scoring* |
| --- | --- | --- | --- | --- | --- | --- | --- | --- |
| ***m.9998T>C*** | **Possibly pathogenic (10 points)** | **Definitely pathogenic (13 points)** | **Pathogenic (PS2, PM10, PM2, PM8)** | **Pathogenic (PS2, PS3, PM2, PM8)** | **Likely pathogenic (PS2, PM2, PP4, BP4)** | **Likely pathogenic (PS2, PS3-supporting, PM2-supporting, PP4, BP4)** | **Likely pathogenic (PS2, PS3-supporting, PM2, PM8)** | **Pathogenic (PS2, PS3, PM2, PM8)** |
| ***m.7530A>G*** | **Possibly pathogenic (10 points)** | **Definitely pathogenic (13 points)** | **Likely pathogenic (PM10, PM7, PM8, PP7)** | **Likely Pathogenic (PS3, PM7, PM8, PP7)** | **VUS (PM2, PP3, PP4)** | **VUS (PS3-supporting, PM2-supporting, PP3, PP4)** | **Likely pathogenic (PS3_supporting, PP3_moderate, PM8, PM2_supporting)** | **Likely pathogenic (PS3, PP3_moderate, PM8, PM2_supporting)** |
| ***m.4305A>G*** | **Possibly pathogenic (10 points)** | **Definitely pathogenic (13 points)** | **VUS (PM10, PM8, PP7)** | **Likely Pathogenic (PS3, PM8, PP7)** | **VUS (PM2, PP4, BP4)** | **VUS (PS3-supporting, PM2-supporting, PP4, BP4)** | **VUS (PS3_supporting, PM8, PM2_supporting)** | **Likely Pathogenic (PS3, PM8, PM2_supporting)** |
| ***m.4271G>C*** | **Possibly pathogenic (7 points)** | **Possibly pathogenic (10 points)** | **Likely pathogenic (PM10, PM8, PP3, PP7)** | **Likely pathogenic (PS3, PM8, PP3, PP7)** | **VUS (PM2, PP3)** | **VUS (PS3-supporting, PM2-supporting, PP3)** | **VUS (PS3_supporting, PM8, PP3, PM2_supporting)** | **Likely pathogenic (PS3_moderate, PM8, PP3, PM2_supporting)** |
| ***m.7453G>A*** | **Definitely pathogenic (15 points)** | **Classification clinvar L/LP**<br>**Variation ID: 869395**<br>**Accession: VCV000869395.2** | **non applicable** | **Pathogenic PS3, PS5, PM2, PP3, PM8, PP7** | **Non applicable** | **Pathogenic PS2, PS3-supporting, PS4-moderate,, PM2-supporting, PP4, PP3** | **non applicable** | **Pathogenic PS3, PM2, PP3, PP6, PP7** |

## 4. Discussion

Systematic whole mtDNA sequencing has increased the detection of pathogenic variants but also the number of VUS, requiring complementary functional analyses for accurate interpretation. To facilitate the implementation of single-fiber analysis in routine diagnostics, we extended the work of Zereg et al. by applying an NGS-based quantification approach. This study provided additional strong evidence supporting the pathogenicity of four novel variants. To our knowledge, the four variants, m.9998T>C in *MT-TG*, m.7530A>G in *MT-TD*, and m.4271G>C and m.4305A>G in *MT-TI*, have not been previously reported in the literature. Using the Yarham’s scoring, all four variants were considered as possibly pathogenic, and applying Wong’s specifications, three of the four were deemed at least likely pathogenic even in the absence of single-fiber analysis, supported by rare population frequency (4/4), tissue segregation (4/4), respiratory chain deficiency (3/4), or pathogenic in silico prediction (2/4). However, under the ClinGen guidelines, three were considered as VUS primarily because this framework does not incorporate tissue segregation data from single patients. Only the m.9998T>C variant was classified as likely pathogenic due to its *de novo* occurrence, which allowed the use of the strong PS2 criterion.

Incorporating single-fiber analysis increased the Yarham classification score by 3 points, upgrading the first three variants m.9998T>C, m.7530A>G, and m.4305A>G, to definitely pathogenic, while m.4271G>C remained possibly pathogenic. Under Wong’s framework, the moderate criterion PM10 was upgraded to the strong criterion PS3, leading to only one change in classification: the m.4305A>G variant progressed from VUS to Likely pathogenic. For the ClinGen classification, PS3 could be applied, however, since no standards exist for objectively analyzing single-fiber data, the Expert Panel recommended using it only as supporting evidence. As a result, single-fiber analysis did not change the ClinGen classification of any of the four variants.

Thus, significant differences can be observed in the application of these guidelines systems. Surprisingly, while single-fiber analysis was previously considered a gold standard in Yarham guidelines, it is not regarded as a strong piece of evidence by the ClinGen classification, providing no upgrade for the three VUS in this study. In fact, the only strong criterion that can be applied for tRNA variants within the ClinGen framework is the *de novo* occurrence criterion (PS2). While this information is often available for children, it is frequently missing in adults. Similarly, the PS4 criterion, which requires more than 16 unrelated probands, is rarely applicable. Furthermore, apart from the *de novo* occurrence, there is no moderate-strength evidence available for tRNA variants, as the PM2 criterion, corresponding to a very low population frequency (Frequency < 0.00002), has been downgraded to PM2-supporting. As in the ACMG classification^8^, at least a moderate level criterion is required to classify a variant as probably pathogenic, it becomes impossible to classify a variant as likely pathogenic if it is not *de novo*, leaving many variants classified as VUS.

Conversely, the “likely pathogenic” status assigned by Wong’s classification appears to be overestimated, with the addition of four moderate-level criteria (PM7 to PM10) and two supporting-level criteria (PP6 and PP7). It should be noted that while the PP6 and PM8 criteria are new, allowing consideration of heteroplasmy levels and tissue segregation, the other criteria correspond to modified weightings of pre-existing ones (PM7 = PP3_moderate, PM9 = PS2_moderate, PM10 = PS3_moderate, PP7 = PM2_supporting).

Our analysis highlights significant differences between the ClinGen and Wong classification frameworks for mitochondrial tRNA variants. While ClinGen appears conservative, often leaving many variants as VUS, Wong’s approach may overestimate the likely pathogenic status due to the weighting of certain criteria such as PM10 and PP6.

These findings suggest that the criteria applied to mitochondrial tRNA variants should be reconsidered. For the ClinGen classification, it may be advisable to assign greater weight to functional analyses using cybrid or single-fiber studies, for example by adopting a PS3-moderate level. It would also likely be beneficial to include a criterion equivalent to PM8 from Wong’s classification, (heteroplasmy (≥5%) across different tissues of an affected individual and its correlation with clinical or biochemical phenotypes), which is an important factor to account for. Regarding Wong’s classification, it appears that the PM10 criterion could be wrongly assigned to non-specific respiratory chain deficiencies or to a few COX-negative fibers that are not significant and may be age-related. Therefore, this criterion should be used very cautiously and potentially downgraded to a supporting-level criterion.

Considering these elements, we chose to use an intermediate classification to interpret the variants identified in this study (Table 2), applying the Wong criteria while downgrading PM10 to a supporting level (Table 3). We also removed the PP6 criterion, as it did not appear sufficiently relevant. Finally, we adapted the names of the new criteria introduced by Wong et al. to align with the original ACMG framework and to remain consistent with other guidelines (PM7 = PP3_moderate, PM9 = PS2_moderate, PM10 = PS3_moderate, PP7 = PM2_supporting).

**Table 3.**
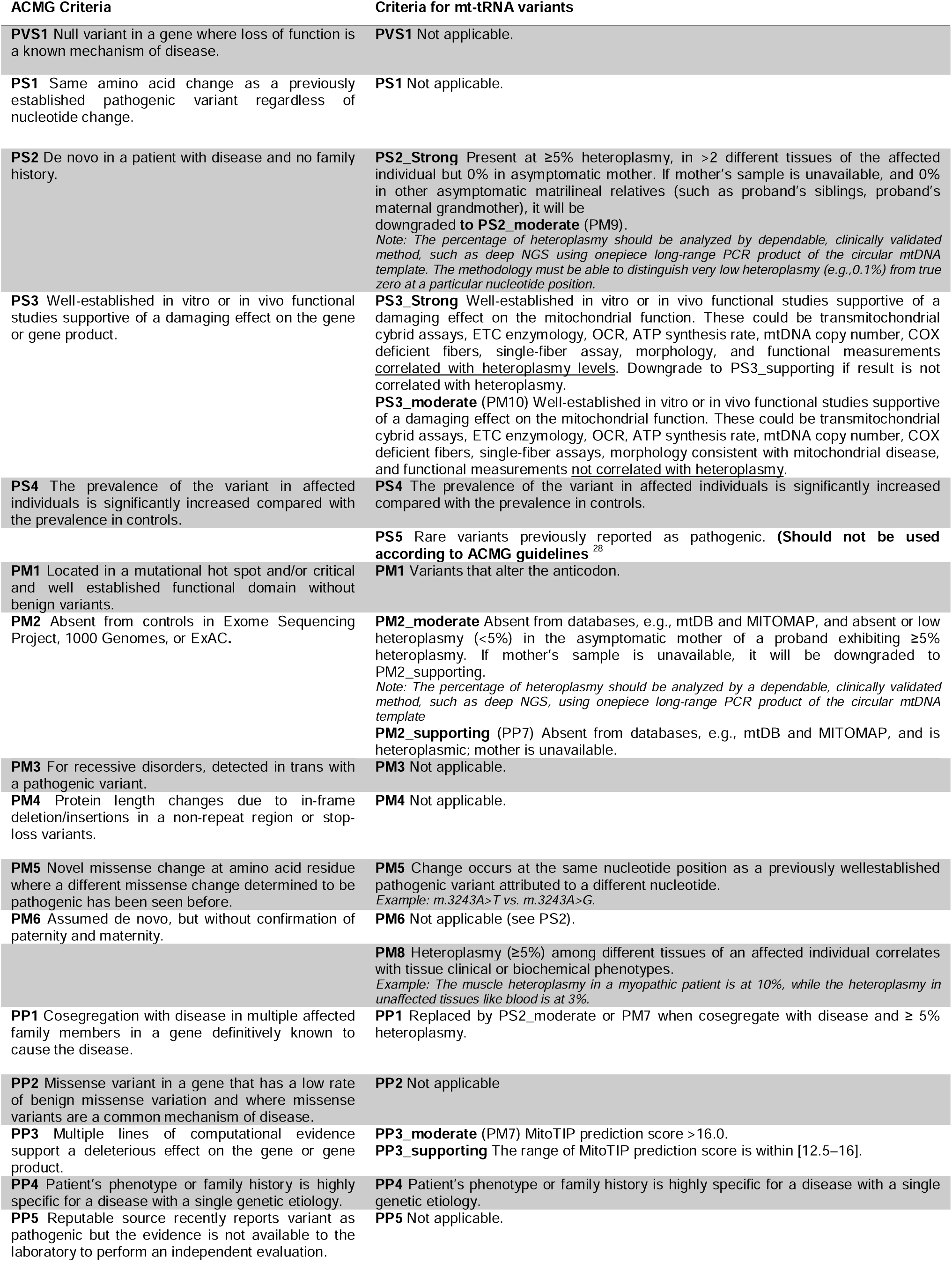
Criteria for the classification of pathogenic mt-tRNA variants.

The m.9998T>C variant affects mt-tRNA^Gly^, encoded by the *MT-TG* gene. Only one variant in this gene has been confirmed as pathogenic in MITOMAP, m.10010T>C. It was reported in an adult-onset mitochondrial myopathy, including muscle weakness, exercise intolerance, myalgia, hyperlactatemia, and combined respiratory chain deficiencies^19^, which is consistent with the clinical features observed in patient 1. Although MitoTIP predicted the m.9998T>C variant as possibly benign, it affects a highly evolutionarily conserved nucleotide position, which is predicted to be involved in tRNA folding (HmtVar) and may consequently disrupt the tertiary structure of mt-tRNA^Gly^^20^. Single-fiber analysis provided additional functional evidence, supporting its reclassification from likely pathogenic to pathogenic.

The m.7530A>G variant affects mt-tRNA^Asp^, encoded by the *MT-TD* gene. Although no variant in this gene has been considered confirmed pathogenic in MITOMAP, several variants have previously been associated with a broad clinical spectrum, ranging from isolated mitochondrial myopathy^21^ to multisystemic mitochondrial disorders including myopathy, spinal ataxia, sensorineural hearing loss, cataract, and cognitive impairment ^22^, as observed in patient 2. This variant alters a highly conserved nucleotide within the D-stem of the tRNA^Asp^ molecule, resulting in an A-to-G substitution that disrupts a canonical Watson–Crick base pair. Consequently, it was predicted as “likely pathogenic” by MitoTIP. Despite these findings and the results from the single-fiber analysis, this variant remains classified as likely pathogenic, mainly due to PM2 (frequency <0.002%) being downgraded to a supporting level.

Both the m.4305A>G and m.4271G>C variants affect the mt-tRNA^Ile^, encoded by the *MT-TI* gene. Numerous variants have been reported in this gene, of which four have been confirmed as pathogenic according to MITOMAP. These variants have been implicated in a wide range of mitochondrial phenotypes, including isolated sensorineural hearing loss, chronic progressive external ophthalmoplegia (CPEO)^23^, Leigh syndrome with severe encephalopathy and progressive cerebral necrosis^24,25^, and familial hypertrophic cardiomyopathy^26^. This phenotypic diversity reflects the clinical heterogeneity associated with mitochondrial tRNA mutations and the frequently limited genotype–phenotype correlation. This is exemplified in our cohort by patients 3 and 4, who exhibited divergent clinical presentations. This clinical difference is likely largely due to the much higher heteroplasmy level in patient 3 compared to patient 4. The m.4271G>C variant resides within a well-characterized post-transcriptional modification site of mt-tRNA^Ile^ ^27^, potentially affecting its secondary structure, stability, or function, and was predicted as likely pathogenic by MitoTIP. In contrast, the m.4305A>G variant was predicted as possibly benign, despite affecting a highly conserved position. Regarding the assignment of PS3, which corresponds to evidence from single-fiber analysis, the m.4271G>C variant was given a moderate strength level (PS3_moderate) due to the limited number of COX-negative fibers analyzed and the high variability among fibers. However, for both variants, the single-fiber analysis enabled their reclassification from VUS to likely pathogenic.

Finally, regarding the m.7453G>A variant in mt-tRNA^Ser^ (*MT-TS1*), it affects a highly conserved nucleotide of the acceptor (ACC) stem, positioned just before the start of the T-stem (Fig.1). This variant has previously been reported as pathogenic in two patients presenting with more severe and earlier-onset phenotypes. It was first identified in a homoplasmic and *de novo* state in a child with fatal neonatal lactic acidosis, associated with severe complex I and IV deficiencies in the heart, skeletal muscle, brain, and liver, as well as a marked depletion of mature tRNA^Ser^ ^14^. It was later reported in a heteroplasmic state in a 15-year-old girl with severe mitochondrial myopathy, who had experienced weakness and fatigue since birth, along with deficiencies in complexes I, III, and IV^13^. Both studies included functional investigations, such as quantification of tRNA^Ser^ (UCN) levels, single-fiber analysis, and cybrid assays. In our cohort, patient 5 presented with a milder phenotype characterized by adult-onset progressive exercise intolerance and CPEO with a lower percentage of COX-negative fibers (30%). Unlike the previously reported cases, which showed very high heteroplasmy levels across all analyzed tissues, patient 5 displayed high heteroplasmy in muscle (96%) but lower levels in urine (43%) and blood (35%), which may explain the isolated muscular involvement and later onset. Single-fiber analysis was consistent with previous findings^13^ and confirmed the contribution of this variant to the patient’s clinical presentation.

Using our adapted criteria, we were able to reclassify the four novel variants before and after functional assessment, providing a more accurate reflection of their clinical relevance. Our study underscores the added value of single-fiber analysis in variant interpretation and supports its integration into routine diagnostic workflows. While NGS has demonstrated strong reliability for heteroplasmy quantification, its high cost and limited scalability remain barriers to widespread application. Future research should focus on developing and validating cost-effective, high-throughput approaches that maintain the diagnostic accuracy required for precise classification.

In conclusion, this study confirmed the molecular diagnosis in five patients and classified four novel variants as pathogenic or likely pathogenic. Although validation on a larger set of known variants is still required, we propose that, for the interpretation of mtDNA tRNA variants, greater weight should be given to tissue segregation and functional analyses compared with the current ClinGen classification. Furthermore, additional work is needed to establish standardized criteria for the objective analysis of single-fiber studies, including the minimum number of fibers, the acceptable proportion of COX-negative fibers, and thresholds for statistical significance. These findings underscore the need to reassess and refine existing guidelines for mtDNA tRNA variant interpretation, integrating functional evidence to provide a more robust and clinically meaningful framework.

## Data Availability

All data produced in the present study are available upon reasonable request to the authors

## ACKNOWLEDGMENTS

I would like to express my special thanks to the laboratory technicians of the Department of Medical Genetics, National Center for Mitochondrial Diseases, research engineers of Inserm U1081, CNRS UMR7284, IRCAN, Nice, France, and Mauri-Crouzet Alessandra for their precious help in carrying out this project.

## CREDIT AUTHORSHIP CONTRIBUTION STATEMENT

**Chloé Prosper**: Formal analysis, Investigation, Writing – Original Draft, Visualization. **Zereg Elamine**: Formal Analysis, Investigation. **Chaussenot Annabelle**: Resources, Writing – Review and Editing. **Bannwarth Sylvie**: Writing – Review and Editing**. Lannes Béatrice, Nathalie Streichenberger, Kaphan Elsa, Nadaj-Pakleza Aleksandra, Villa Luisa, Sacconi Sabrina, Masingue Marion:** Resources. **Francou Bruno:** Data curation, Writing – Review and Editing. **Saadi Ait-El-Mkadem Samira:** Data curation, Writing – Review and Editing. **Fragaki Konstantina**: Data curation, Writing – Review and Editing. **Paquis-Flucklinger Véronique**: Writing – Review and Editing. **Rouzier Cécile**: Conceptualization, Methodology, Validation, Formal analysis, Writing – Review and Editing, Supervision, Project administration.

---

**Table S1.**
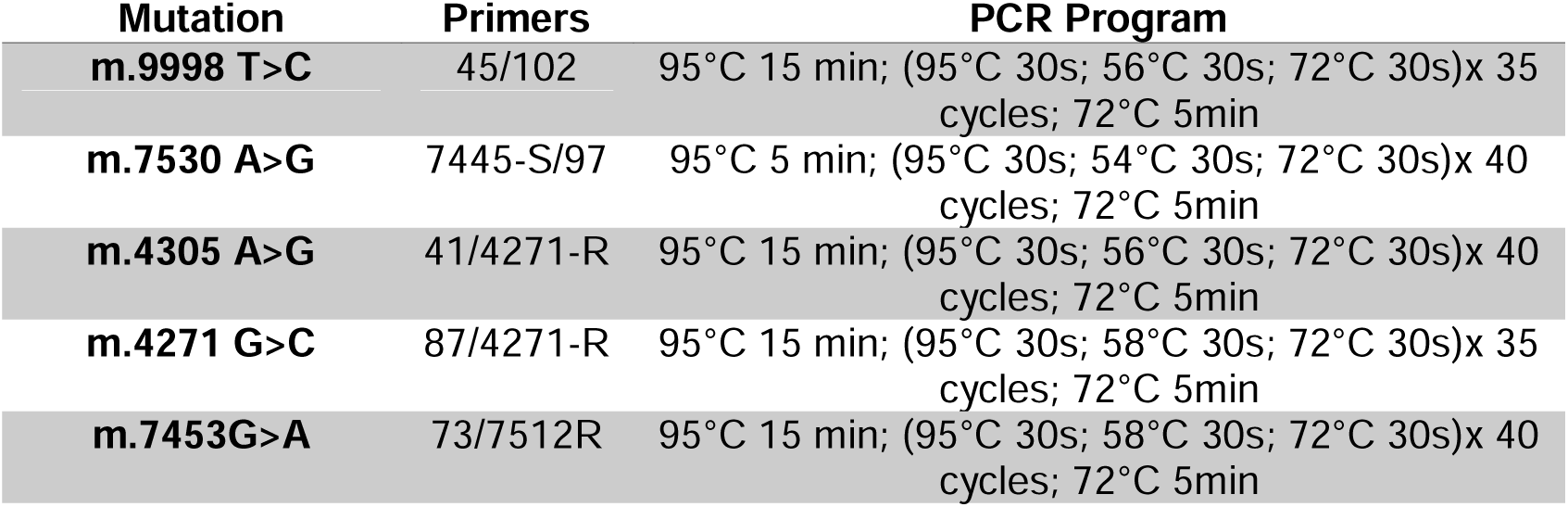
Primers used in PCR reactions for the five mt-DNA variants.

**Table S2.**
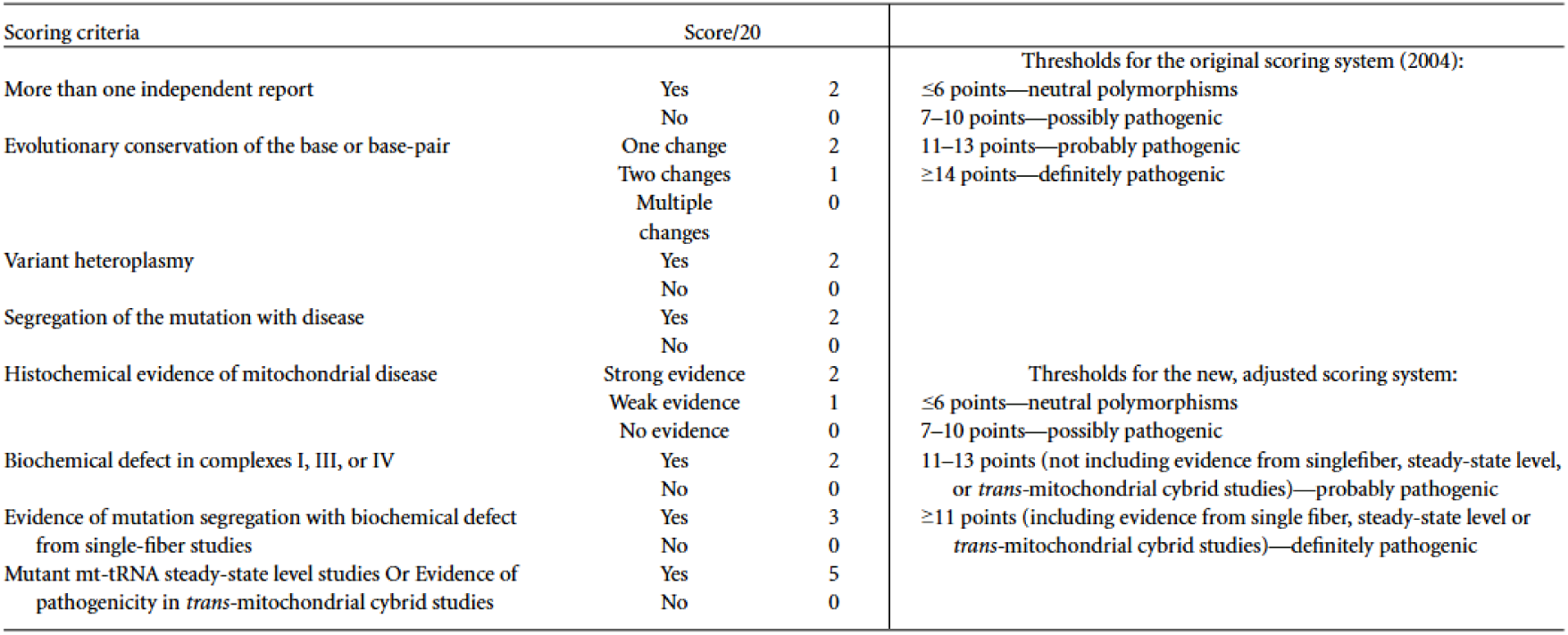
Scoring system establish by Yarham et al. 2011.

